# State-wide health systems strengthening initiatives for systematic tuberculosis passive case finding under programmatic settings in Haryana-Early implementation findings

**DOI:** 10.1101/2025.10.21.25338493

**Authors:** Hitesh Verma, Kathirvel Soundappan, Akash Ranjan Singh, Prabhadevi Ravichandran, Kedar Mehta, Suseendar Shanmugasundaram, Sandhya Gupta, Kuldeep Singh, Hemant Deepak Shewade

**Author notes:** **Address for correspondence** Dr Hitesh Verma, State TB Officer, State TB cell, Office of DGHS, Sector 6, Panchkula, Haryana, India, 9467003812. **Role of investigators** **1. HV:** Principal Investigator and corresponding author, conception / design of the protocol, acquisition of data, data analysis / interpretation, drafting / critically reviewing the paper, giving approval for the final version to be published **2. KS, ARS, PR, SG, HDS, KS, KM, SS:** conception/design of the protocol, data analysis/interpretation, critically reviewing the paper, for approving the final version to be published.

## Abstract

**SETTINGS:** Systematic tuberculosis (TB) passive case finding initiative in Haryana, a high-TB burden state in north India that has an annual shortfall of presumptive TB examinations.

**OBJECTIVE:** To compare the changes in enrolling persons with presumptive TB in public-sector peripheral health institutions (PHIs) between July–September (before) and October–December 2024 (during initiative).

**DESIGN:** In this quasi-experimental (before and during) study design, a multi-pronged health systems strengthening initiative was implemented state-wide: i) enrollment of PHIs below primary health centre level in *Ni-kshay* (web-based TB information management system), ii) identification of a nodal person from each PHI-without TB diagnosis centre (TDC) for enrolling presumptive TB in *Ni-kshay* and iii) identification of community volunteers for transport of sputum to TDCs.

**RESULTS:** The enrolment of over 10 presumptive TB at PHIs with TDC remained at 87%, while it increased from 51% to 63% in non-TDC PHIs. The mean number of sputum transport increased from 12 to 22 per non-TDC PHI. The presumptive TB enrolment rate increased from 1020 to 1660 per 100,000 population.

**CONCLUSION:** Early implementation findings are encouraging. We intend to sustain this in 2025 with additional interventions, culminating in early diagnosis and reducing the missing TB.

## INTRODUCTION

Globally in 2023, an estimated 10.6 million people developed tuberculosis (TB), of them 3.1 million were missed.^1^ The identification of all persons with TB and linking them to care is essential to reduce the incidence and mortality due to TB, which are the targets of the End TB strategy.^2^

India shares the highest burden of TB with 26% (2.8 million) of estimated annual global TB incidence and 28% (0.3 million) of estimated global deaths.^1^ The National Tuberculosis Elimination Program (NTEP) in India has implemented various strategies to identify all people with TB and link them with care.^3^ Active and passive case finding are the two broad strategies adopted by the program to identify all people with TB. However, both strategies, if implemented simultaneously, may prove much more effective than otherwise. Active case finding (ACF) potentially helps in detecting patients with poor awareness and related healthcare-seeking behaviour, especially in vulnerable and marginalised communities.^4^ However, this consumes heavy resources. In converse, passive case finding (PCF) is a person-initiated pathway that utilizes routine health systems to identify persons with presumptive TB. Here, the health systems must be adequately oriented and equipped with resources and built-in capacities to identify and follow up on all people with presumptive TB.^4^

The program routinely collects and reports presumptive TB examination rates against the given target at various levels since it is directly linked with the TB notification rate. In India, PCF is done only for 30% of the people with presumptive TB coming to the hospital.^5^ So, there is a scope for improvement in TB screening through strengthening the PCF. Further, 91% to 97% of the people with chest symptoms who visited the healthcare provider (any) missed or were not screened with sputum microscopy, which indicates an important missed opportunity in identifying potential people with TB.^6^ Hence, there is an urgent need to strengthen the PCF at all peripheral health institution (PHI) levels using the strategies and tools available within the program.^3^

Haryana is a state with the second highest TB prevalence (477 per 100,000 population) after Delhi in India.^7^ The annual TB notification in the state is 261 per 100,000 population, but it notified only 84% of the expected people with TB in 2023.^4^ To further improve this, multi-pronged health systems strengthening initiative was implemented statewide to improve the TB diagnostic care cascade (from identification of people with presumptive TB to testing and diagnosis) in public sector PHIs with and without a TB diagnosis centre (TDC). The focus was on primary healthcare level public PHIs without a TDC (non-TDC PHI). Within one quarter of implementation, this paper reports the early implementation findings of changes in presumptive TB enrolment, stratified by PHIs with and without a TDC compared to before implementation.

## METHODS

### Study design

This was a quasi-experimental (before-during the health systems strengthening initiative) study involving routinely collected program data.

### Study population

All PCF-detected persons with presumptive TB enrolled (irrespective of examination) from July-September 2024 (before) and October-December 2024 (during) in all public PHIs of Haryana. ACF-detected and household contact tracing-detected persons with presumptive TB were excluded. The data was accessed from *Ni-kshay* Portal on January 20, 2025

### Study setting

#### General setting

As per the Census 2011, the total population of the Haryana state is 25.4 million with 22 districts, 140 community development blocks and 7356 villages.^8^ The healthcare delivery system has 22 district hospitals, 50 sub-district hospitals, 128 community health centres, 514 primary health centres (PHC) and 2727 health subcentres.^9^ NTEP is operational in all districts of Haryana. The presumptive TB enrollment rate of the state is 1643 (national average of 1710) per 100,000 population in 2023.^4.^ The TB notification rate in the state (261 per 100,000 population) is higher than the national average (178.8 per 100,000 population).^10^

#### Specific setting

The District TB Centre (DTC) is the nodal point for TB under NTEP. At the block/sub-district level, Tuberculosis Units (TUs, n=173) implement NTEP activities, with one TU for approximately 200,000 population. Each TU has a Senior Treatment Supervisor (STS) and Senior TB Laboratory Supervisor, supervised by a Medical Officer-TB Control. TUs oversee PHIs with or without TDC. All PHIs provide TB treatment services. In 2024, there were 3336 public PHIs, of which 444 were operational TDC during July to September and 476 were operational TDC during October to December. Of 3141 public primary healthcare level PHIs, 213 were operational TDCs. Public primary healthcare level PHIs include PHC with a medical doctor that covers ≈30000 population and health subcentres with auxiliary nurse midwives (ANMs) and/or community health officers (CHOs) covering ≈5000 population.

Healthcare providers, including ANMs, CHOs, and Medical Officers, identify persons with presumptive TB via symptom screening. Persons with presumptive TB are referred to TDCs for diagnosis after enrollment in *Ni-kshay*, a web-based TB information management system of NTEP. Laboratory technicians record test results in *Ni-kshay*, and positive cases are initiated on treatment as per program guidelines, with STS ensuring follow-up. Thus, enrolling persons with presumptive TB formally initiates the TB care cascade, making it a crucial step.

### Health systems strengthening initiative

From October 2024, to strengthen the TB enrollment mechanism which potentially may lead to improvement in the PCF, a multi-pronged intervention was implemented state wide (details in Box 1); The three core components of the intervention area, i) increasing the enrollment of health subcentres and *AYUSH* (Ayurveda, Yunani, Siddha and Homeopathy) private clinics in *Ni-kshay*, ii) identifying a nodal person for identifying and enrolling (in *Ni-kshay*) presumptive TB in public non-TDC PHIs, and iii) identification and engagement of community volunteers and fixing their honorariums for sputum collection and transport to TDCs (₹ 50 from PHI and ₹ 100 from residence). This is for the person with presumptive TB enrolled in *Ni-kshay* but not undergone a sputum examination.

### Data variables, sources of data and data collection

A total of seven key performance indicators for non-TDC PHIs were identified and clubbed into three groups: i) training and sensitization indicators, which included two indicators, ii) screening and sputum transport-related indicators, which included three indicators, and iii) enrollment indicators, which included two indicators. The purpose of grouping these indicators was to help programme managers at the state and district level to routinely monitor the functioning of PHIs in terms of enrolment. This was included in the routine monitoring and evaluation framework of NTEP in the state. (TABLE 1)

**Table 1:** Performance indicators before and during the initiative of systematic improvement of TB enrolment in routine programme setting in Haryana, India 2024.

| Sr No | Indicators for systematic passive case finding initiative | Before (July-Sep 2024)<br>(N=2892) | After(Oct-Dec 2024)<br>(N=2860)<sup>#</sup> |
| --- | --- | --- | --- |
|  | **Training and sensitization indicators** |  |  |
|  | **At PHI level** |  |  |
|  | 1. Proportion of PHIs with identification of staff* and job actualization for marking “P” in outpatient register for persons with presumptive TB and enrolling in *Ni-kshay* | 1721 (59.5%) | 2221 (77.7%) |
|  | 2. Proportion of PHIs having staff trained for sample collection and triple packing of the samples obtained from the person with presumptive TB attending outpatient department | 1721 (59.5%) | 2221 (77.7%) |
|  | **Screening and sputum transport indicators** |  |  |
|  | **At PHI level** |  |  |
|  | 3. Proportion of PHIs enrolled in *Ni-kshay* portal | 1606 (56.2%) | 2021 (70.7%) |
|  | 4. Proportion of PHIs with identification and linking of community volunteers to transport samples to TDC | 438 (15.1%) | 1374 (48%) |
|  | 5. Number of samples transported per PHIs by courier service provider/community volunteer to TDC | 11.6 | 22.4 |
|  | **Enrolment indicators** |  |  |
|  | **At the state level** |  |  |
|  | 6. Proportion of identified people<sup>#</sup> with presumptive TB offered TB test (Microscopy/NAAT)** | 18915(38.4%) | 58562(52.6%) |
|  | **At PHI level** |  |  |
|  | 7. Proportion of PHIs enrolling at least one presumptive TB | 1675 (57.9%) | 2011 (70.3%) |

Footnote: #A total 32 PHIs without TDC have been upgraded to TDC after September 2024; \*Drug-resistant TB coordinator at the district level and Senior treatment laboratory supervisor at sub district level has been assigned for the implementation of initiative to systematic improvement of TB enrolment in routine programme setting; # Non-TDC identified 49258 and 111334 people with presumptive TB, ‘before’ and ‘during’ intervention period. \*\* Information obtained from *Ni-kshay* portal, PHI-peripheral health institute; TDC-TB diagnostic centre, NAAT: Nucleic acid amplification test

Data sources include the *Ni-kshay* enrollment register for patient enrollment and demographic details and the NeoGeo site (Haryana’s digital geospatial healthcare directory, detailing population distribution and facilities available) for population data of PHIs. TB type and notification were extracted from the *Ni-kshay* presumptive case notification and laboratory registers. Operational definitions used in the study are mentioned in BOX 2.

### Data management and statistical analysis

The PHI enrollment register, presumptive case laboratory register and notification register of *Nikshay* were merged to create a single data frame using the *Ni-kshay* ID as a unique linking variable. Any person enrolled twice in two or more PHIs had their IDs deduplicated. Data was analysed using the R software package and R Studio Version 1.3.1093© 2009-2020 RStudio, PBC. The codes of analysis were saved in the R markdown file (Supplementary material 1). Descriptive analysis for the seven key performance indicators in PHIs without a TDC were represented as frequencies and percentages (TABLE 1). A difference in change analysis was conducted to assess the early implementation effect of the health systems strengthening intervention on presumptive TB enrollment across PHIs with and without TDC before and during the initiative.

### Ethics approval

Ethical approval was obtained from the Institutional Human Ethics Committee of the ICMRNational Institute of Epidemiology, Chennai, India (NIE/IHEC/A/202408-02 dated August 7, 2024). The study was conducted after the necessary administrative approvals from the Health Department of Haryana. The health systems strengthening initiative are being implemented by the study state as a quality improvement exercise to improve the routine NTEP programmatic outcomes. As the study involved the extraction of routinely captured secondary data, a waiver for informed consent was sought and obtained from the ethics committee.

## RESULTS

Of the total 614,393 enrolled people with presumptive TB, 286,032 were PCF-detected during the study period. Of them, 56% were offered microscopy or nucleic acid amplification test (NAAT) to confirm TB disease. Of the total presumptive TB (n=286,032), 9.3% were notified as TB disease. Of the total notified, 64% were bacteriologically confirmed. The treatment initiation rate was 93% among the bacteriologically confirmed and 98% clinically diagnosed TB. (FIGURE 1)

**Figure 1:**
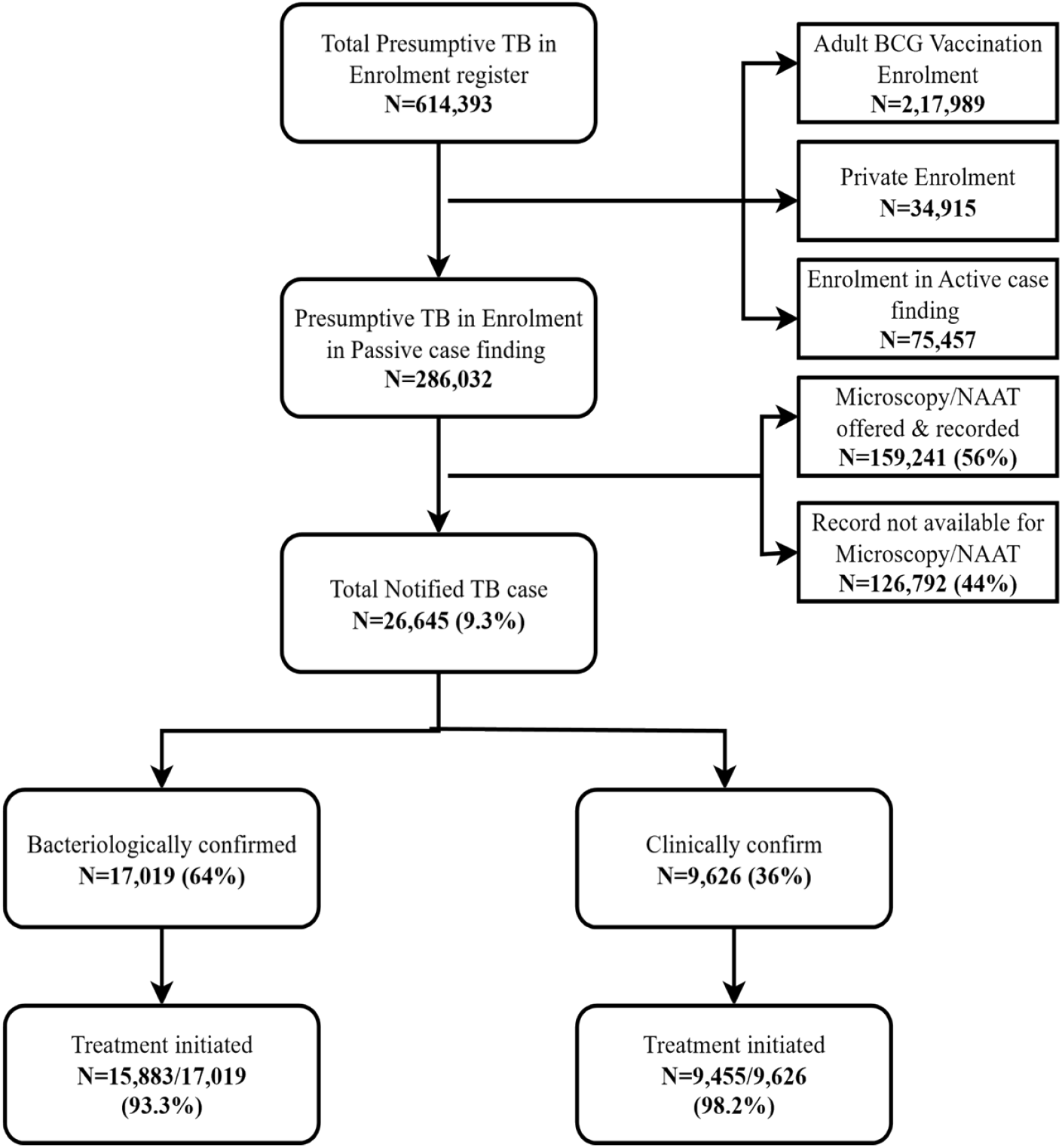
Cascade of care of total presumptive TB cases identified at Public Health Institutions of Haryana (India) from July-December 2024. Footnote: BCG-Bacillus Calmette-Guerin; NAAT: Nucleic Acid Amplification test; it includes CBNAAT and TrueNAT.

Under the training and sensitization domain, the proportion of non-TDC PHIs with identification and job actualization for marking “P” in the outpatient register and enrolling into *Ni-kshay* increased from 59.5% to 77.7%. A similar increase in the availability of staff trained in the collection and packing of sputum samples was observed.

Under the screening and sputum transport domain, the proportion of non-TDC PHIs out of the total PHIs enrolled in *Ni-kshay* increased from 56.2% to 70.7%. Similarly, there was an improvement in the identification of community volunteers. The number of sputum specimens transported by courier service providers or community volunteers increased from 12 to 22 per PHIs without a TB diagnosis centre per quarter (TABLE 1).

Under enrollment indicators, the proportion of presumptive TB identified from non-TDC PHIs who received tests for microbiological confirmation increased from 38.4% to 52.6%. The proportion enrolling at least one person with presumptive TB also increased. (TABLE 1)

During October-December 2024 (during intervention), the proportion of TDC-PHI enrolling >10 persons with presumptive TB remained unchanged at 87%, while it increased to 63% (from 51% in July-September 2024) among non-TDC PHIs. (FIGURE 2)

**Figure 2:**
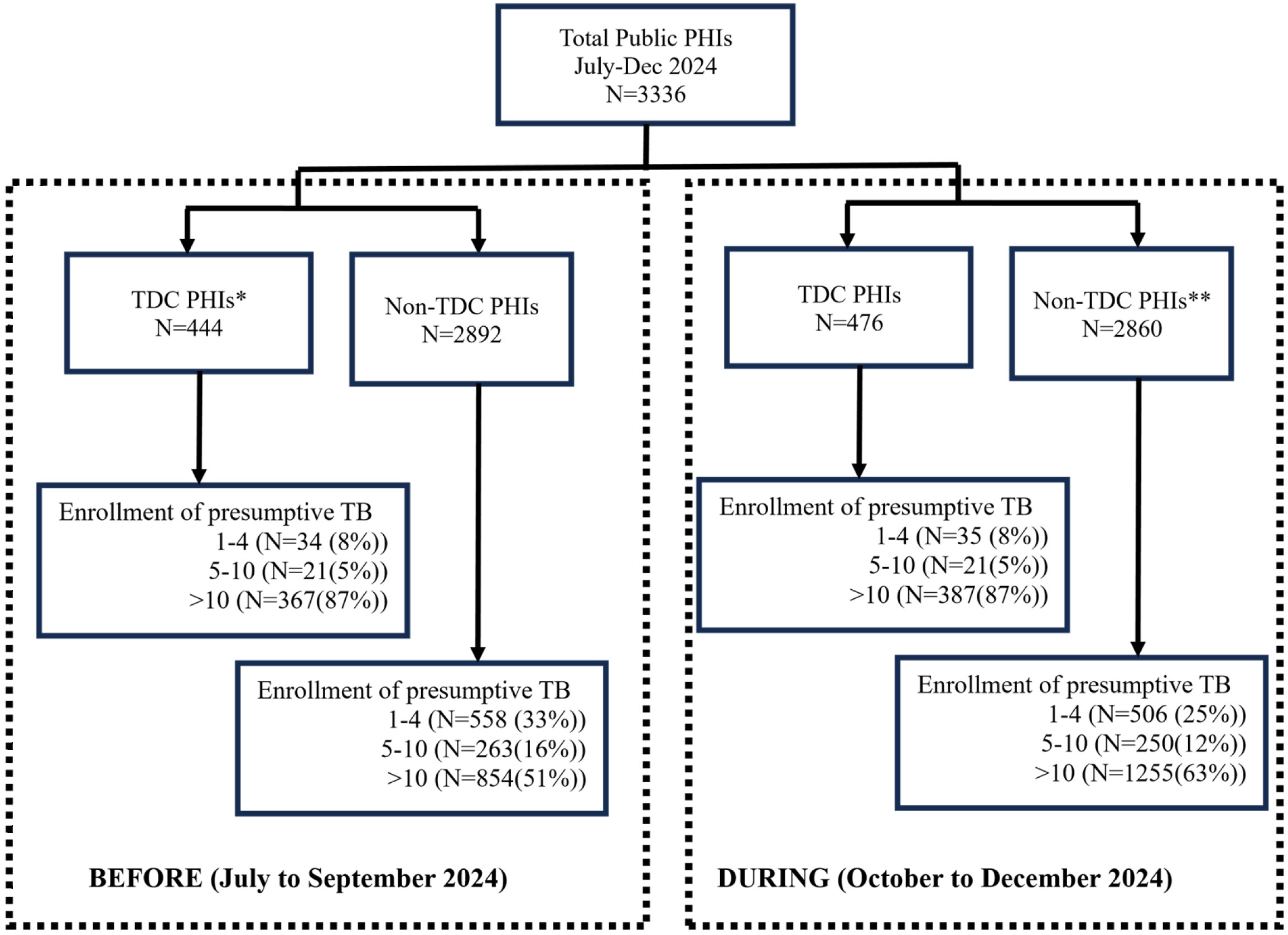
Enrolment of people with presumptive TB among PHIs with and without a TB diagnostic centre in Haryana before initiative (July to September 2024) and during initiative 401 (October to December 2024) Footnote: *Total Number of TDC-PHIs enrolling presumptive TB were 422 and 443 before after intervention respectively. **Total Number of non-TDC PHIs enrolling presumptive TB were 1675 and 2011 before after intervention respectively

Presumptive TB enrollment per PHI per quarter (non-annualized) has increased by 14.7% (177 to 203) in TDC-PHI and by 129.4% (17 to 39) in non-TDC PHI. (FIGURE 3) Hence, 114.7% increase in non-TDC PHI can be attributed to the initiative. The overall presumptive TB enrolment rate increased from 4082 to 6642 per 100,000 population. The quarterly presumptive TB enrollment rate (non-annualized) per 100,000 population at the state level increased from 1020 (before) to 1660 (during initiative).

**Figure 3:**
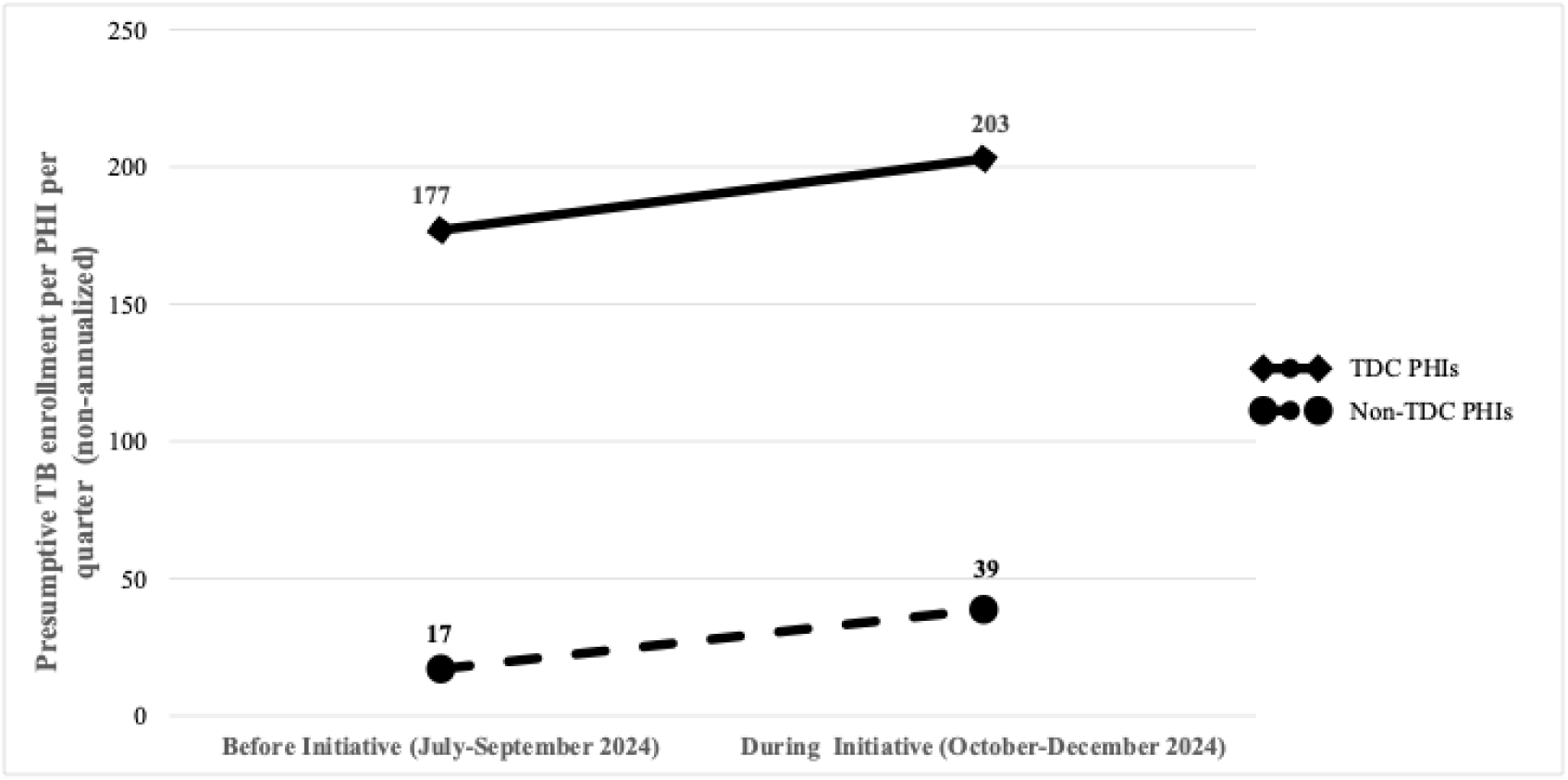
The change of presumptive TB enrollment per PHI with and without TDC Haryana, India (Jul-Sep 2024 vs. Oct-Dec 2024) Footnote: PHI-peripheral health institute; TDC-TB diagnostic centre

## DISCUSSION

This paper is probably the first published study reporting the effect of a multi-pronged health systems intervention at the state level to improve TB PCF in India. The initiative in Haryana, a high-TB burden state in northern India, led to improved screening practices in outpatient departments, especially in non-TDC PHIs. This resulted in an increase in the number of nonTDC PHIs enrolling people with presumptive TB and specimens transported from PHI or patients’ homes to the TDC. This eventually resulted in improved presumptive TB enrollment.

However, only half of the enrolled presumptive TB cases underwent microscopy or NAAT testing.

This state-wide study has huge potential to improve PCF. This initiative, along with key performance indicators that focus on non-TDC PHIs, may be replicated in other states. These indicators are robust and sensitive enough for the performance assessment of PHIs for passive case detection. However, the program managers must be flexible and dynamic in choosing the health systems strengthening interventions according to local contexts that have the potential to improve the TB diagnostic care cascade in PCF settings. Studies have reported that strategies for ACF can improve the yield of detection, but strengthening of PCF that too in a programmatic setting has been little discussed.^11,12^ Within less than a quarter of a year, the initiative doubled the presumptive TB enrollment in non-TDC PHIs and increased quarterly presumptive enrollment at the state level. Identification, sensitization and job actualization of existing staff may augment PCF. As observed in this study, it should be backed with efficient specimen transport systems.^13^ The state TB programme engaged the national postal service to transport the specimens, as it was already used during the COVID-19 pandemic period.^14^ To make this even more flexible, the community volunteers were also engaged for this purpose.^15^ This initiative puts the study state with an added advantage as the state has recently (January 2025) rolled out the upfront NAAT testing (3-4 sites per district) for all presumptive TB.

Lemaire et al. tested a multi-pronged intervention using a before-and-after study design to improve the TB PCF and treatment outcome among the less than 15 years age group in nine sub-Saharan African countries.^16^ The intervention bundle included a) symptom-based screening at entry points of health facilities by health workers, b) capacity building of health workers on TB prevention, diagnosis and treatment, c) sample collection and transportation, including expansion of the NAAT services, and d) household contact investigation. This study observed a 46% increase in monthly paediatric TB case detection during the intervention period, especially a 17% increase in cases notified from the lower-level health facilities.^16^ Similar to our study, the proportion of children with presumptive TB undergoing confirmatory TB tests was lower (22.8%) in the later study, which was attributed to the non-consideration of TB as a differential diagnosis by the clinicians. Similarly, Wandwalo et al. tested a bundle of quality improvement interventions in PHIs to improve the TB PCF in Tanzania, and observed a 52% increase in the TB case notification after intervention.^17^

There was a gap in bacteriological confirmation before and during the initiative (only half of the presumptive TB cases underwent microscopy or NAAT). This highlights the gap in diagnostic and reporting mechanisms. Anecdotal experience suggested that for many people with presumptive TB whose examination result is negative, laboratory technicians do not prioritize data entry into *Nikshay*. In addition to improving the data entry, there is a need to identify those who do not get tested due to suboptimal laboratory capacity. A bundled intervention (diagnostic-fluoresence microscopy and NAAT and on-site training of non-physician health workers) in cross-border African settings showed a rapid and 24 times higher chance for bacteriological confirmation.^18^ Beyond sputum collection and transportation, the recent rollout of upfront NAAT in the study state has the high potential to improve the proportion of people with presumptive TB offered bacteriological confirmation tests. The effect of this added health systems strengthening may be assessed in future operational research.

The limitations are that these are early implementation effects. Sustenance of improved presumptive TB enrolment (overall and in non-TDC PHIs) needs to be studied in 2025, along with the eventual effect on improving TB notification and early diagnosis. Initial efforts put in an initiative are hard to sustain, especially for initiatives requiring multiple steps to achieve an outcome. Such initiatives often have reduced effectiveness due to challenges later in the care cascade.^19,20^

## CONCLUSION

The state-wide, multi-pronged health systems strengthening initiative for systematic TB passive case finding in Haryana has shown promising results. Within a quarter, the initiative successfully engaged the non-TDC PHIs, improving outpatient screening practices and sputum transportation, resulting in the doubling of presumptive enrolments in these PHIs and an increase in enrollments at the state level. However, a critical gap was that only half of the presumptive TB cases underwent confirmatory testing. These findings emphasize the need for stronger diagnostic linkages, robust data management systems, and continuous monitoring of these PHIs over long term, eventually improving TB detection and reducing diagnosis delay.

## Data Availability

The data underlying the results presented in the study are available with authors

## Box, Tables and Figures

### Box 1: Details of health systems strengthening initiative for the systematic passive case finding in Haryana with focus on PHIs without a TB diagnosis centre

1. Sensitization of the DTOs and providing training to field-level staff in data management was done.
2. All identified persons with presumptive TB were marked as “P” in the OPD register. The P-marked patients were enrolled in *Ni-kshay* the same day by the nodal staff.
3. The state signed a BNPL (Buy now pay later) MoU with India Post to transport sputum samples.
4. Either of the three mechanisms was used to collect sputum samples
  i. The patient deposits the sputum sample in the lab of the PHIs without a TDC and the same is transported via a government human carrier
  ii. The sputum sample is collected from home by a private human carrier and transported to the nearest TDC (Rs 100/-per sputum sample was sanctioned),
  iii. The sputum sample is collected in the PHIs without a TDC, packed and transported to the nearest TDC by India Post.
5. The nodal staff was trained and accounted for the collection and triple packing of sputum samples before their transportation through the India Post/Courier as per NTEP guidelines.
6. The TB test results (both positive and negative) were documented in the Ni-kshay portal and the laboratory register at TDC.
7. MOTCs did a daily review of STLS at TU.
8. DTO did a weekly review of MOTCs.
9. Fortnightly review meetings to ensure proper implementation of the strategies.

*Footnote: DTO: District Tuberculosis Officer, OPD-outpatient department, MoU: Memorandum of understanding, PHI-peripheral health institute; TDC-TB diagnostic centre; NTEP-National TB Elimination Program; MOTC-Medical Officer-TB Control; STLS-Senior TB Laboratory Supervisor; STO-State TB Officer*.

### Box 2: Operational Definitions

1. People with Presumptive Pulmonary TB refers to a persons with any of the symptoms and signs suggestive of pulmonary TB, including cough for ≥two weeks, fever for ≥two weeks, significant weight loss, haemoptysis, or any abnormality in the chest radiograph. The following are also to be investigated as presumptive Pulmonary TB
  a. Contacts of Microbiologically confirmed TB patients having cough of any duration
  b. Presumptive /confirmed extra-pulmonary TB having cough of any duration
  c. HIV positive patient having cough of any duration
2. Presumptive extra-pulmonary TB refers to the presence of organ-specific symptoms and signs like swelling of lymph nodes, pain and swelling in joints, neck stiffness, disorientation etc., and/or constitutional symptoms like significant weight loss, persistent fever for ≥two weeks, or night sweats.
3. Presumptive Paediatric TB refers to children with persistent fever and/ or cough for ≥two weeks, loss of weight*/ no weight gain and/ or history of contact with infectious TB cases** *History of unexplained weight loss or no weight gain in the past three months; loss of weight is defined as loss of more than 5% body weight as compared to the highest weight recorded in the last three months. ** In a symptomatic child, contact with a person with any form of active TB within the last two years may be significant
4. Passive case finding (PCF): It refers to a patient-initiated pathway to TB diagnosis when the patient voluntarily reports symptoms to the Medical Officer. Intensified case finding is a component of PCF that involves systematic screening of all people seeking care in a health facility or a clinic for identification of people
5. Indicators:
  a. Presumptive TB examination rates (PTBER) is defined as the number of people with presumptive TB tested per 100,000 population
  b. Proportion of presumptive TB offered molecular diagnostics upfront for diagnosis of TB: Of the presumptive TB tested, the proportion that was offered a rapid molecular test for diagnosis of TB as the first test of diagnosis.
  c. Annualised TB case notification rate (ACNR) is defined as the number of TB cases notified per lakh population on an annualised basis.

## References

1. World Health Organization. Global tuberculosis report, 2024. Geneva, Switzerland: WHO, 2024.

2. World Health Organization. The End TB Strategy, 2015. Geneva, Switzerland: WHO, 2015.

3. Khanna A, Saha R, Ahmad N. National TB elimination programme - What has changed. Indian J Med Microbiol. 2022;42:103–107

4. Ministry of Health and Family Welfare, Government of India. India TB Report 2024: New Delhi, India: Central TB Division, Directorate General of Health Services, 2024.

5. Pala S, Bhattacharya H, Lynrah K, Sarkar A, Boro P, Medhi G. Loss to follow up during diagnosis of presumptive pulmonary tuberculosis at a tertiary care hospital. J Family Med Prim Care. 2018;7(5)942.

6. Subbaraman R, Nathavitharana RR, Satyanarayana S, et al. The Tuberculosis Cascade of Care in India’s Public Sector: A Systematic Review and Meta-analysis. PLoS Med. 2016;13(10).

7. Central TB Division, Ministry of Health and Family Welfare, Government of India. National TB Prevalence Survey India, 2019–2021. New Delhi, India, 2023

8. Directorate of Census Operations Haryana. Census. Ministry of Home Affairs, Government of Haryana. 2011. Accessed August 2, 2024. https://haryana.census.gov.in/census

9. Health Department Haryana. Haryana Health Infrastructure. 2024. Accessed August 2, 2024. https://haryanahealth.gov.in/infrastructure/

10. Ministry of Health and Family Welfare, Government of India. Leading the way: India TB Report 2023: New Delhi, India: Central TB Division, Directorate General of Health Services, 2023.

11. Shewade HD, Gupta V, Ghule VH, et al. Impact of Advocacy, Communication, Social Mobilization and Active Case Finding on TB Notification in Jharkhand, India. J Epidemiol Glob Health. 2019;9(4)233–242.

12. Kranzer K, Afnan-Holmes H, Tomlin K, et al. The benefits to communities and individuals of screening for active tuberculosis disease: a systematic review [State of the art series. Case finding/screening. Number 2 in the series]. International Journal of Tuberculosis and Lung Disease. 2013;17(4)432–446.

13. Dama E, Nikiema A, Nichols K, et al. Designing and Piloting a Specimen Transport System in Burkina Faso. Health Secur. 2020;18(S1)S98–S104.

14. Yadav M, Jain AK, Singhal R, Chadha M, Arora VK, Bhargava A. Incidence and Patterns of Drug Resistance in Patients with Spinal Tuberculosis: a Prospective, Single-Center Study from a Tuberculosis-Endemic Country.Indian J Orthop. 2023;57(11)1833–1841.

15. Garg K, Panwar A, Sharma N, et al. Integrating Indian Post and National Tuberculosis Elimination Program - A new way ahead. Journal of Comprehensive Health. 2021;9(2)94–96.

16. Lemaire JF, Cohn J, Kakayeva S, et al. Improving TB detection among children in routine clinical care through intensified case finding in facility-based child health entry points and decentralized management: A before-and-after study in Nine Sub-Saharan African Countries. PLOS Global Public Health. 2024;4(2)e0002865.1-19

17. Wandwalo E, Kamara D V, Yassin MA, et al. Enhancing Tuberculosis Case-Finding: A Case of Quality Improvement Initiative in Tanzania. Trop Med Infect Dis. 2022;7(6)97:1-7

18. Manabe YC, Zawedde-Muyanja S, Burnett SM, et al. Rapid Improvement in Passive Tuberculosis Case Detection and Tuberculosis Treatment Outcomes After Implementation of a Bundled Laboratory Diagnostic and On-Site Training Intervention Targeting Mid-Level Providers. Open Forum Infect Dis. 2015;2(1)1–10

19. Chambers DA, Glasgow RE, Stange KC. The dynamic sustainability framework: Addressing the paradox of sustainment amid ongoing change. Implementation Science. 2013;8(1)1–11.

20. Pai M, Schumacher SG, Abimbola S. Surrogate endpoints in global health research: still searching for killer apps and silver bullets? BMJ Glob Health. 2018;3755.

